# Effectiveness of 13-valent pneumococcal conjugate vaccine against medically-attended lower respiratory tract infection and pneumonia among older adults

**DOI:** 10.1101/2021.06.30.21259721

**Authors:** Joseph A. Lewnard, Katia J. Bruxvoort, Heidi Fischer, Vennis X. Hong, Lindsay R. Grant, Luis Jódar, Alejandro Cané, Bradford D. Gessner, Sara Y. Tartof

**Author notes:** Corresponding author: Joseph Lewnard, 2121 Berkeley Way, Room 5410, Berkeley, California 94720.

## Abstract

**Importance:** In 2014, the US Advisory Committee on Immunization Practices (ACIP) extended existing pneumococcal vaccination recommendations for adults aged ≥65 years to include 13-valent pneumococcal conjugate vaccine (PCV13), primarily to prevent non-bacteremic pneumonia.

**Objective:** To determine PCV13 effectiveness against all-cause inpatient plus outpatient medically-attended lower-respiratory tract infection (LRTI) and pneumonia among US older adults.

**Design:** Prospective, open cohort study following participants from 2016 to 2019. We conducted analyses in a self-matched framework, comparing outcomes during participants’ follow-up periods before and after receipt of PCV13.

**Setting:** Kaiser Permanente Southern California (KPSC) integrated healthcare delivery system.

**Participants:** Adults aged ≥65 years who received PCV13 between 2016-2019.

**Exposures:** Receipt of PCV13 at ages ≥65 years, concordant with ACIP guidelines.

**Main outcomes and measures:** We estimated the adjusted hazards ratio (aHR) for first LRTI and pneumonia episodes during each respiratory season, comparing PCV13-exposed and PCV13-unexposed time at risk for each participant using a self-matched inference framework. We computed aHR estimates using Cox proportional hazards models. We defined vaccine effectiveness (VE) as (1–aHR)×100%. We also estimated PCV13-attributable absolute reductions in incidence of LRTI and pneumonia.

**Results:** Observations were available both before and after PCV13 receipt for 42,700 participants. Among these individuals, 1,419 experienced LRTI and 969 experienced pneumonia over approximately 26,000 combined years of follow-up before PCV13 receipt; 3,849 experienced LRTI and 2,727 experienced pneumonia over approximately 74,000 combined years of follow-up after PCV13 receipt. In adjusted analyses, VE was 9.5% (95% confidence interval: 2.2% to 16.3%) against all-cause medically-attended LRTI and 8.8% (–0.2% to 17.0%) against all-cause medically-attended pneumonia. In contrast, we did not identify evidence of protection against LRTI and pneumonia following receipt of 23-valent pneumococcal polysaccharide vaccine. We estimated that PCV13 prevented 0.7 (0.2 to 1.4) and 0.5 (0.0 to 1.0) cases of LRTI and pneumonia, respectively, per 100 vaccinated persons annually. Over a five-year time horizon, one case of LRTI and pneumonia, respectively, was prevented for every 27 (14 to 116) and 42 (–97 to 268) individuals receiving PCV13.

**Conclusions and relevance:** PCV13 vaccination among older adults reduced the burden of medically-attended respiratory illness in this population.

**KEY POINTS:** *Question:* Does 13-valent pneumococcal vaccine (PCV13) reduce the burden of lower respiratory-tract infection (LRTI) and pneumonia among older adults?

*Findings:* In this longitudinal cohort study of adults aged 65 years and older from 2016-2019, PCV13 reduced incidence of LRTI and pneumonia episodes due to any cause by 9.5% and 8.8%, respectively. This effect corresponded to prevention of 0.7 and 0.5 episodes of LRTI and pneumonia, respectively, per 100 vaccinated persons each year. Protection from 23-valent pneumococcal polysaccharide vaccine was not apparent.

*Meaning:* Among older adults, PCV13 vaccine administration substantially reduces the burden of medically-attended LRTI and pneumonia.

## INTRODUCTION

In the United States, implementation of 13-valent pneumococcal conjugate vaccine (PCV13) in the pediatric immunization schedule has reduced the incidence of pneumococcal and all-cause pneumonia among older adults.^1^ Despite this, and decades of direct vaccination of older adults with 23-valent pneumococcal polysaccharide vaccine (PPSV23), PCV13-targeted serotypes continue to account for substantial disease burden, including 49% of pneumococcal pneumonia cases with a serotype documented and 28% of invasive pneumococcal disease (IPD) cases among US adults.^2,3^

Following a randomized, placebo-controlled trial in The Netherlands demonstrating PCV13 efficacy against non-bacteremic pneumonia attributable to PCV13-serotype pneumococci among older adults,^4^ the US Advisory Committee on Immunization Practices (ACIP) recommended in 2014 that—in addition to an existing recommendation for PPSV23 to prevent invasive pneumococcal disease (IPD)—all adults aged ≥65 years should receive PCV13. This recommendation aimed primarily to prevent non-bacteremic pneumonia^5^, which accounts for the majority of pneumococcal disease cases, deaths, and healthcare costs among US adults.^6^ However, the burden of disease prevented by use of PCVs among older adults remains poorly understood.^7^ In light of this uncertainty, ACIP revised pneumococcal vaccination guidelines in 2019 to state that adults aged ≥65 years without high-risk conditions should receive PCV13 based on shared clinical decision-making with their healthcare providers.^8^ As other countries have not issued similar recommendations for healthy older adults to receive conjugate vaccination with PPSV23, assessing the public health benefits of PCV13 use among US older adults is a priority to inform vaccination strategies in other settings, and to guide implementation of next-generation conjugate vaccines (PCV15/20) targeting additional serotypes.

We estimated effects of PCV13 administration among adults aged ≥65 years on risk of all-cause LRTI and pneumonia. We used findings from these analyses to quantify the incidence of LRTI and pneumonia prevented by PCV13 direct effects among older adults.

## METHODS

### Setting

The study cohort comprised members of Kaiser Permamente Southern California (KPSC) health plans. The KPSC healthcare system provides comprehensive, integrated care to approximately 19% of the population of southern California. Members are enrolled through employer-provided, pre-paid, and federally-sponsored insurance plans, and closely resemble the demographic composition of the geographic area.^9^ Care delivery (including diagnoses, immunizations, prescriptions, and services provided) across outpatient, emergency department, hospital, and virtual care settings is captured through electronic health records (EHRs), while care received out-of-network is captured through insurance claims, yielding near-complete ascertainment of healthcare interactions for KPSC members.

The protocol for this study was reviewed and approved by the KPSC Institutional Review Board.

### Population and follow-up

The analytic cohort included individuals aged ≥65 years as of 1 September of the years 2016, 2017, 2018, and 2019, who had been enrolled in KPSC health plans for ≥1 year previously (to capture prior healthcare utilization and comorbidities). Individuals were eligible for inclusion if they received PCV13 at any point from 1 September, 2016 to 31 August, 2019, so that data were available from at least one respiratory disease season (beginning 1 September of each year) before and after PCV13 receipt for each individual. We excluded individuals receiving PCV13 or PPSV23 in sequences that were not concordant with ACIP guidelines at ages ≥65 years (defined as: receipt of PPSV23 ≥1 year after PCV13 for immunocompetent individuals or ≥8 weeks after PCV13 for immunocompromised individuals; or receipt of PCV13 ≥1 year after PPSV23). We included data from the 2016-17 season onward to avoid bias introduced by transition from use of the 9^th^ to 10^th^ revisions of International Classification of Diseases, clinical modification (ICD-9-CM/ICD-10-CM) coding systems during the 2015-16 season.^10^

We defined observations for each individual, for each year, from 1 September to 31 August of the ensuing calendar year; we truncated observations in 2019 at 31 December to exclude effects of the coronavirus disease 2019 (COVID-19) pandemic. Because KPSC uses automated alerts prompting healthcare providers to offer pneumococcal vaccination to individuals with high risk of pneumonia, including individuals who recently had LRTI or pneumonia diagnoses, we truncated observations each year at the first occurrence of a study endpoint (LRTI or pneumonia, as defined below), or at dates of receipt of PCV13 or PPSV23. To allow for time to onset of immunity, we also excluded person-time contributed over the first 30 days after receipt of PCV13 or PPSV23.

### Outcomes

Study endpoints were the first diagnosed, medically-attended (i.e., inpatient or outpatient [including emergency department]) LRTI episode or pneumonia episode during each year of follow-up, defined according to ICD-10-CM codes (**Table S1**). We censored observations each year at death, disenrollment, or mid-season receipt of the study vaccinations (PCV13, PPSV23, or zoster vaccination), as described below.

### Exposures

The primary exposure of interest was ACIP guideline-concordant receipt of PCV13 at ages ≥65 years. We also recorded dates of receipt of other recommended adult vaccines, including PPSV23, seasonal influenza vaccines (live or inactivated), and zoster vaccines (live or recombinant). We considered seasonal influenza vaccines and zoster vaccines as positive and negative control exposures, respectively, for assessment of bias, as confounding factors in the relationship between pneumococcal vaccination and respiratory disease outcomes could apply similarly to these vaccines.^11^ Whereas seasonal influenza vaccination was expected to reduce individuals’ risk of study endpoints,^12^ zoster vaccination was not expected to impact these disease outcomes.^13^

We further recorded individuals’ age at the beginning of each year, their history of healthcare utilization in the prior year (including numbers of outpatient visits, emergency department visits, and inpatient admissions), sex, race/ethnicity, tobacco smoking history, body mass index, history of comorbid conditions, and median household income within their residential census tract. Prior year comorbid conditions were determined from ICD-10-CM codes (**Table S2**). We further recorded whether individuals met ACIP definitions of “high risk” or “at risk” status for pneumococcal vaccine prioritization (**Table S3**) based on medical history.^8^

## Statistical analysis

We estimated vaccine effectiveness (VE) using a self-matched inference framework comparing outcomes during periods before and after receipt of vaccination from the same individual. This analytic approach was selected to mitigate confounding driven by notable differences in demographics and risk factor prevalence within the full cohort of KPSC cohort members aged ≥65 years who received or did not receive PCV13 (**Table S4**). For our primary analyses of PCV13 effectiveness, we estimated the adjusted hazards ratio (aHR) for first LRTI and pneumonia diagnoses using Cox proportional hazards models accounting for individuals’ age, receipt of seasonal influenza vaccination (defined as a time-varying exposure), and presence of the risk factors described above. We defined VE=(1-aHR)×100%. We computed robust standard errors using the sandwich estimator to correct for repeated observations from each individual. Models accounted for unique baseline hazards defined within strata for each individual according to their history of receipt of adult vaccinations other than PCV13 (PPSV23 and zoster vaccination) at the beginning of each year; thus, PCV13 VE estimates address differences in risk for the same individual, before and after PCV13 receipt, holding other vaccine exposures constant. Within each season, we modeled seasonal influenza vaccination as a time-varying exposure via the Andersen-Gill framework.^14^

While this first-event analysis framework could mitigate bias resulting from pneumonia episodes serving as a catalyst for PCV13 receipt, seasonal differences in the likelihood of a respiratory infection being due to pneumococcus could lead to differences in VE estimates based on the start date defining a year. Consequently, we conducted sensitivity analyses defining the start of each year as 1 July or 1 November.

We used the same approach for secondary analyses addressing VE for PPSV23, seasonal influenza vaccines, and zoster vaccines. Here, we defined strata for individuals’ history of receipt of PCV13 and zoster vaccines (for analyses of PPSV23 VE) and receipt of PCV13 and PPSV23 (for analyses of zoster VE). To estimate VE for seasonal influenza vaccines, we defined strata for individuals’ history of receipt of PCV13, PPSV23, and zoster vaccines. All analyses treated seasonal influenza vaccination as a time-varying exposure within seasons.

Last, we estimated the incidence of medically-attended LRTI and pneumonia averted by direct effects of PCV13 among older adults within the full KPSC cohort aged ≥65 years. Defining observed incidence among PCV13-vaccinated individuals as *λ*_1_, we estimated the counterfactual incidence among vaccinated individuals, without PCV13, as *λ*_0_ = *λ*_1_/(1 − VE), and the incidence of disease averted by PCV13 as *λ*_0_ − *λ*_1_. Based on evidence that PCV13-conferred protection persists ≥5 years among older adults,^4^ we defined the number of older adults requiring vaccination to prevent one case, at one-year and five-year time horizons, as 1/(*λ*_0_ − *λ*_1_) and 1/[5(*λ*_0_ − *λ*_1_)], respectively. We calculated *λ*_1_ over the seasons with full 12-month observation periods (2016-17, 2017-18, and 2018-19). We considered rates for incidence of any LRTI or pneumonia each year (allowing, at maximum, one episode per individual each season), consistent with our VE estimation framework, and for all LRTI or pneumonia episodes each year not preceded by a prior episode within 30 days.

## RESULTS

### Cohort characteristics and risk factors

In total, 42,700 individuals met inclusion criteria for our analytic cohort (**Table 1**) among 673,677 individuals aged ≥65 years who were enrolled in KPSC health plans for ≥1 year between 1 September, 2016 and 31 December, 2019; exclusions included 117,072 (17.4% of 673,677) individuals who did not receive PCV13 by 31 December, 2019, 371,869 (55.2%) individuals who received PCV13 before 1 September, 2016, and 142,036 (21.1%) individuals who did not have successive years of follow-up before and after PCV13 receipt during the study period (due to death or disenrollment following PCV13 receipt, or receipt of PCV13 after 1 September, 2019).

**Table 1:** Outcomes among the analytic cohort over the study period.

| Year | Vaccination status, during follow-up period | Total population | Observed outcomes<sup>1</sup> |  |
| --- | --- | --- | --- | --- |
|  |  |  | Any LRTI during follow-up | Any pneumonia during follow-up |
| 2016-17 |  |  |  |  |
|  | Before PCV13 receipt | 39,572 | 942 | 660 |
| 2017-18 |  |  |  |  |
|  | Before PCV13 receipt | 12,201 | 469 | 295 |
|  | After PCV13 receipt | 29,449 | 1736 | 1,100 |
|  | Total | 41,650 | 2,205 | 1,395 |
| 2018-19 |  |  |  |  |
|  | Before PCV13 receipt | 3,162 | 68 | 56 |
|  | After PCV13 receipt | 38,102 | 1,846 | 1,402 |
|  | Total | 41,264 | 1,914 | 1,458 |
| 2019<sup>2</sup> |  |  |  |  |
|  | After PCV13 receipt | 38,950 | 679 | 521 |
| Total period |  |  |  |  |
|  | Before PCV13 receipt | 42,700 | 1,419 | 969 |
|  | After PCV13 receipt | 42,700 | 3,849 | 2,727 |
|  | Total | 42,700 | 4,954 | 3,472 |

1. Counts indicate the number of unique individuals with any LRTI or pneumonia diagnosis during the period indicated, rather than the total number of episodes.
2. Monitoring in 2019 ended 31 December 2019.

Overall, 47.9% (20,442/42,700) of individuals within the analytic cohort were aged 65-69 years, 48.5% (20,719/42,700) were non-Hispanic whites, and 55.9% (23,888/42,700) belonged to the ACIP “normal risk” stratum for pneumococcal disease (**Table 2**). Overall, within the full KPSC cohort, individuals who received PCV13 at ages ≥65 years were older than non-recipients, had higher rates of healthcare utilization than non-recipients, and higher prevalence of each chronic comorbid condition (**Table S4**). These differences between the analytic cohort and the general KPSC membership were less pronounced (**Table S5**). Differences in demographic attributes and risk factor prevalence between PCV13 recipients and non-recipients were mirrored in comparisons of recipients and non-recipients of PPSV23, zoster vaccination, and seasonal influenza vaccines (**Tables S6**-**S7**).

**Table 2:**
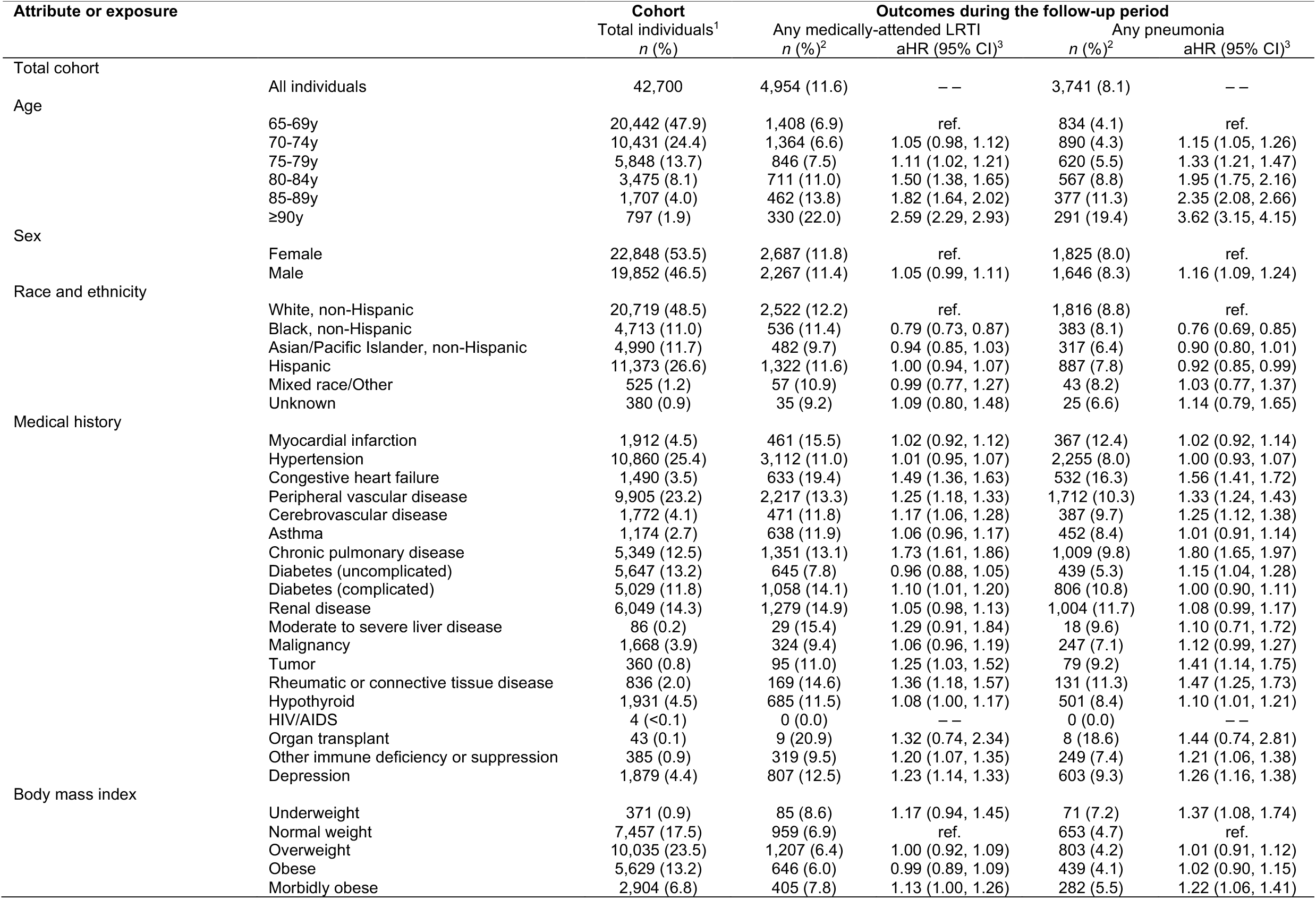
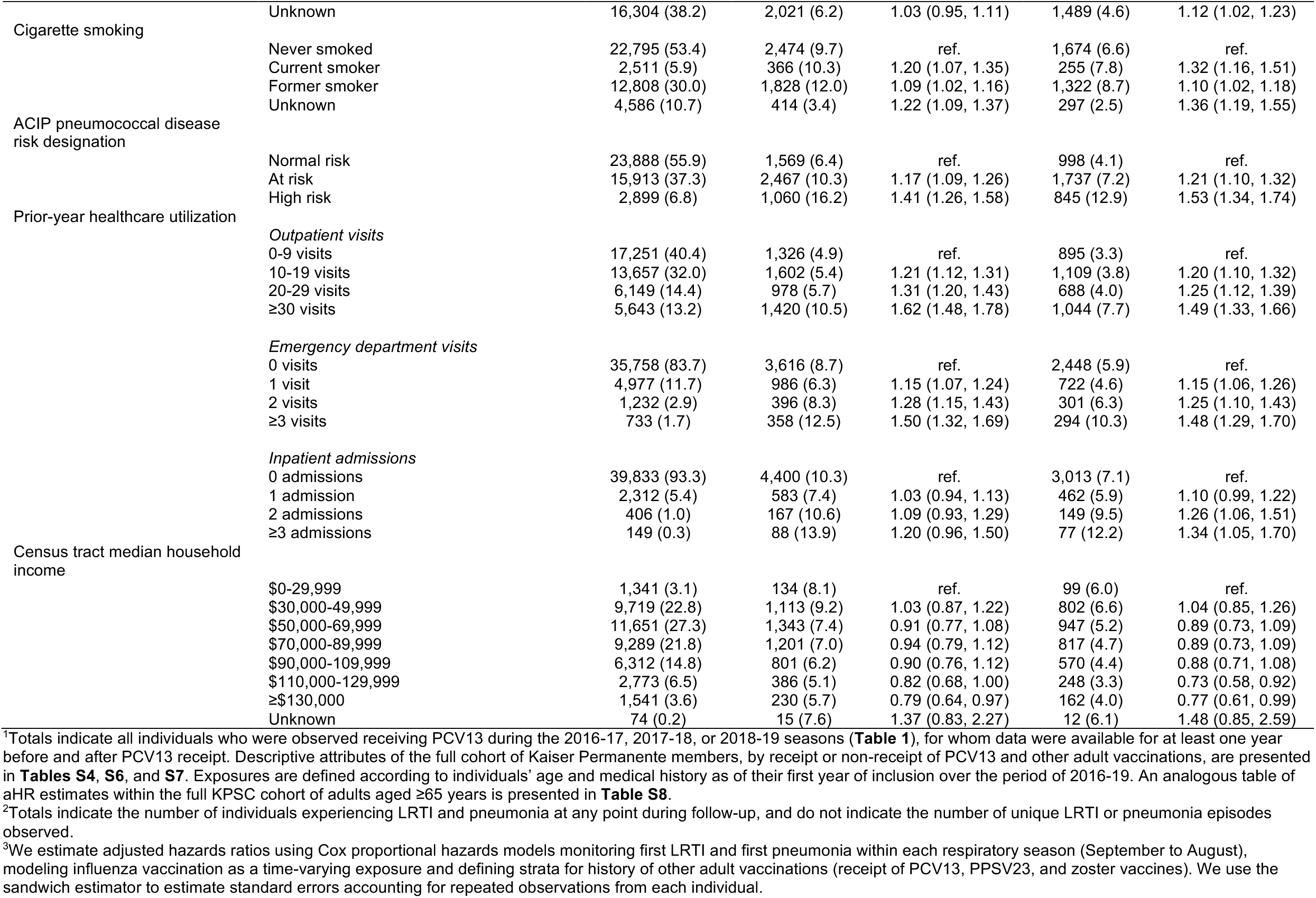
Attributes of the analytic cohort and associated risk of medically-attended lower respiratory tract infection and pneumonia due to any cause.

Within the analytic cohort, increased risk of LRTI and pneumonia was independently associated with older age, male sex, smoking, morbid obesity, and history of medical conditions, most notably congestive heart failure, peripheral vascular disease, chronic pulmonary disease, moderate-to-severe liver disease, immune deficiency/immunosuppression, and depression (**Table 2**). Belonging to ACIP “at risk” or “high risk” strata was independently associated with higher risk for both outcomes. Higher rates of outpatient, emergency department, and inpatient care interactions in the prior year were each independently associated with greater risk of LRTI and pneumonia. Risk for both outcomes was modestly lower among individuals residing in higher-income census tracts. These findings were mirrored in analyses of the full cohort of KPSC members aged ≥65 years (**Table S8**).

### Vaccine effectiveness

Within the analytic cohort, incidence rates of first LRTI and pneumonia were 5.7 and 3.8 episodes per 100 person-years, respectively, during periods preceding PCV13 receipt, and 5.6 and 4.0 episodes per 100 person-years during periods after PCV13 receipt (**Table 3**; **Table S9**; **Figure 1**). When comparing rates during the years immediately before and after PCV13 receipt within this cohort, point estimates of incidence rates for both LRTI and pneumonia were lower in the year after vaccination than the year before vaccination in 17 of 25 age strata (**Figure 1**). Overall, we estimated 9.5% (2.2-16.3%) VE against first LRTI in self-matched analyses controlling for age, within-season influenza vaccination, and comorbid conditions (**Table 3**). Under the same analysis approach, we estimated 8.8% (–0.2-17.0%) VE against first episodes of pneumonia.

**Table 3:**
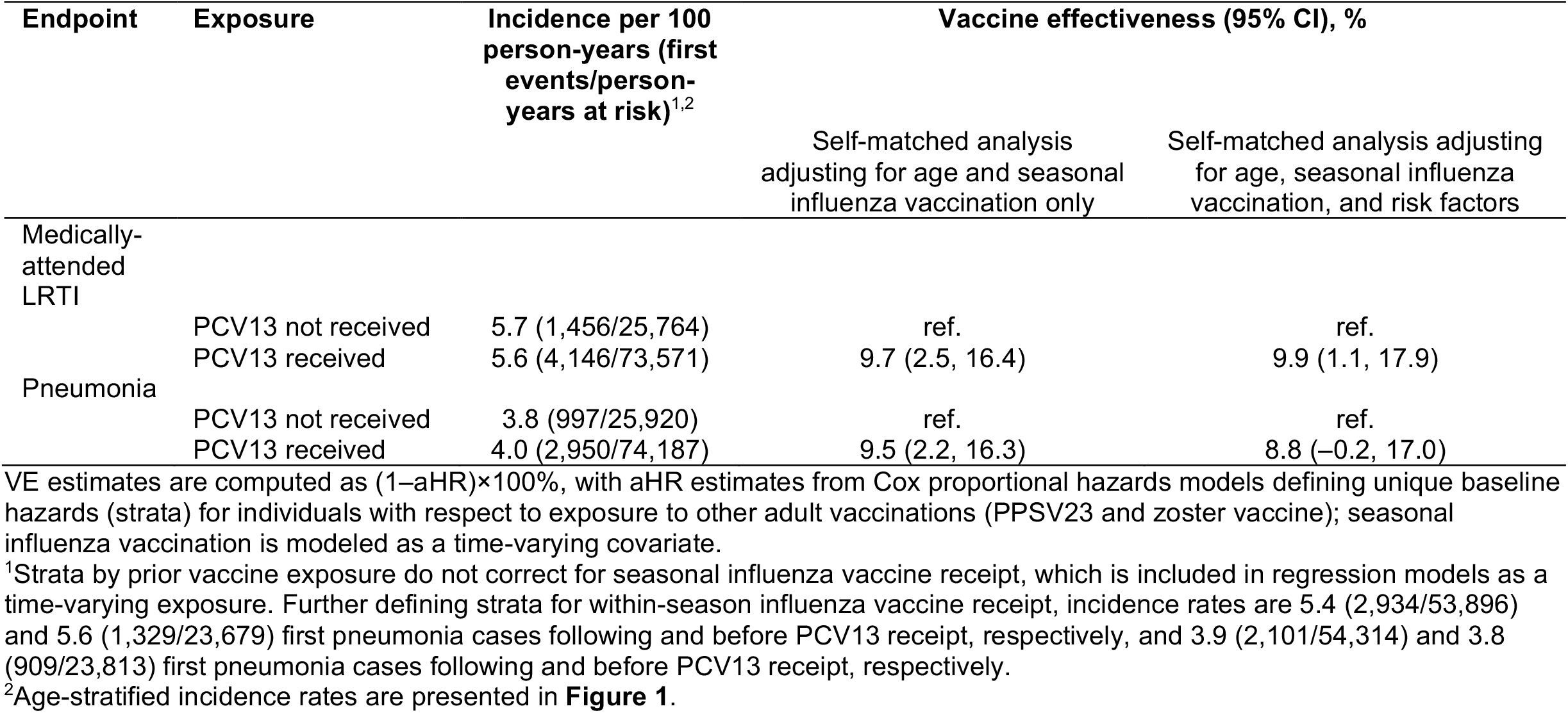
Effectiveness of PCV13 against medically-attended lower respiratory tract infection and pneumonia due to any cause.

| Endpoint | Exposure | Incidence per 100 person-years (first events/person-years at risk)<sup>1,2</sup> | Vaccine effectiveness (95% CI), % |  |
| --- | --- | --- | --- | --- |
|  |  |  | Self-matched analysis adjusting for age and seasonal influenza vaccination only | Self-matched analysis adjusting for age, seasonal influenza vaccination, and risk factors |
| Medically-attended LRTI | PCV13 not received | 5.7 (1,456/25,764) | ref. | ref. |
|  | PCV13 received | 5.6 (4,146/73,571) | 9.7 (2.5, 16.4) | 9.9 (1.1, 17.9) |
| Pneumonia | PCV13 not received | 3.8 (997/25,920) | ref. | ref. |
|  | PCV13 received | 4.0 (2,950/74,187) | 9.5 (2.2, 16.3) | 8.8 (−0.2, 17.0) |

VE estimates are computed as $(1 - \text{aHR}) \times 100\%$ , with aHR estimates from Cox proportional hazards models defining unique baseline hazards (strata) for individuals with respect to exposure to other adult vaccinations (PPSV23 and zoster vaccine); seasonal influenza vaccination is modeled as a time-varying covariate.
<sup>1</sup>Strata by prior vaccine exposure do not correct for seasonal influenza vaccine receipt, which is included in regression models as a time-varying exposure. Further defining strata for within-season influenza vaccine receipt, incidence rates are 5.4 (2,934/53,896) and 5.6 (1,329/23,679) first pneumonia cases following and before PCV13 receipt, respectively, and 3.9 (2,101/54,314) and 3.8 (909/23,813) first pneumonia cases following and before PCV13 receipt, respectively.
<sup>2</sup>Age-stratified incidence rates are presented in **Figure 1**.

**Figure 1:**
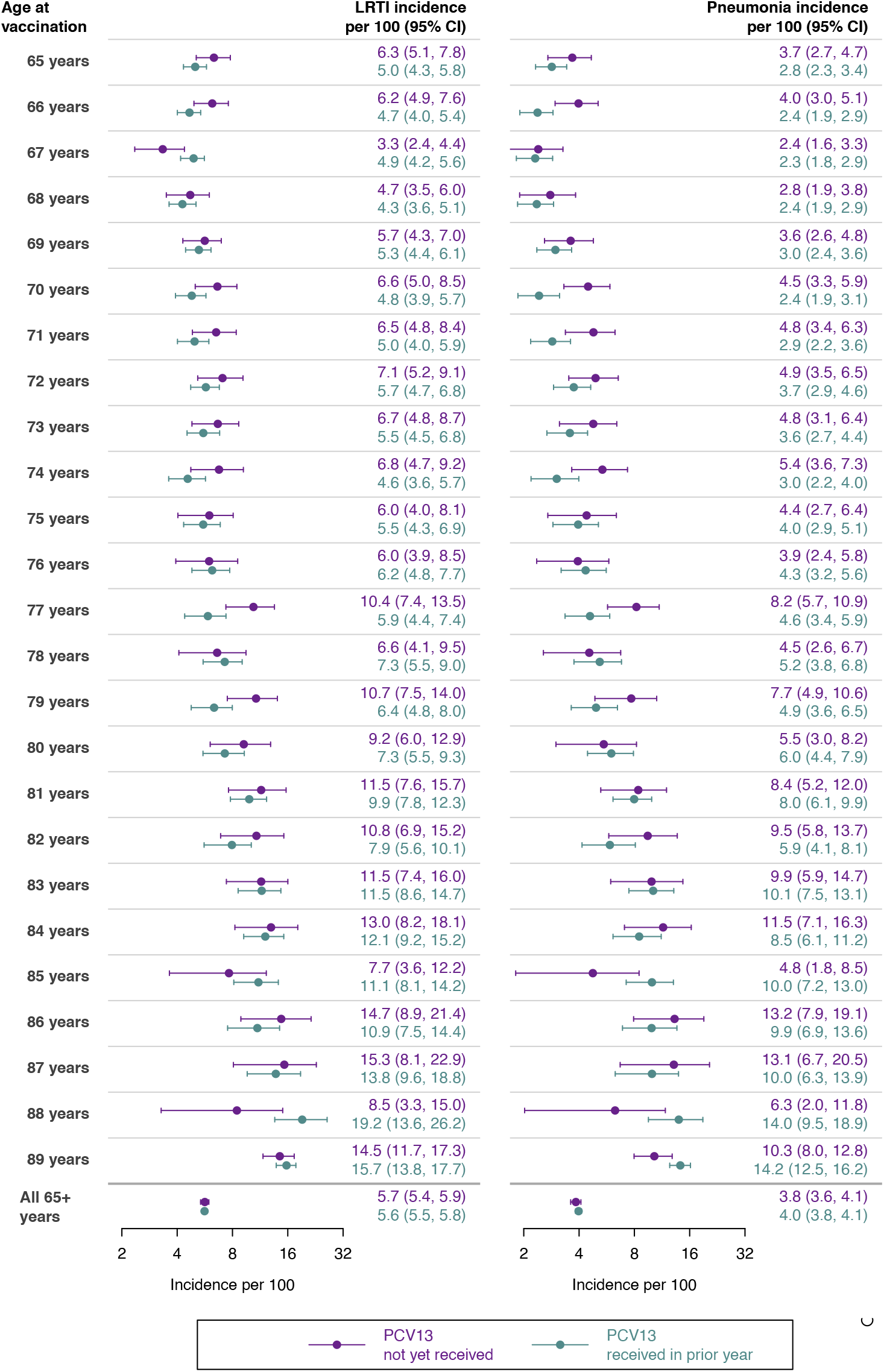
Incidence of first LRTI and pneumonia during years before and after receipt of PCV13. We present incidence rates for LRTI (left) and pneumonia (right) during the seasons immediately preceding and following receipt of PCV13, stratified by age at receipt of PCV13. We obtain 95% credible intervals via bootstrap resampling.

We obtained marginally higher VE estimates in sensitivity analyses monitoring first LRTI and pneumonia episodes over seasons defined as 1 November-31 October, and marginally lower estimates when defining seasons from 1 July-30 June, consistent with the winter peak in pneumococcal disease risk^15^ (**Table S10**). In secondary analyses, we did not identify evidence of PPSV23 effectiveness against either LRTI or pneumonia (**Table S11**). For seasonal influenza vaccination, we estimated 15.1-17.6% VE against first LRTI and 16.3-18.0% VE against first pneumonia, consistent with high VE reported for seasonal influenza vaccines against confirmed influenza A and B infections over the study period (**Table S12**). Zoster vaccination, which we considered a negative-control exposure, was not associated with protection against either LRTI or pneumonia (**Table S11**).

We estimated that PCV13 averted 0.7 (0.2-1.4) LRTI and 0.5 (0.0-1.0) pneumonia episodes annually, respectively, for every 100 adults aged ≥65 years receiving vaccination (**Table 4**). Over a single year of vaccine protection, 1 LRTI episode would be prevented for every 134 (71-580) individuals vaccinated, while over five years, 1 LRTI episode would be prevented for every 27 (14-116) individuals vaccinated; correspondingly, the number needed to vaccinate to prevent 1 pneumonia case was 210 (–484-1,341) over one year and 42 (–97-268) over five years. Among 460,750 cohort members who had received PCV13 by the start of the 2018-19 season, we estimated that PCV13 prevented 2,822 (552-5,195) and 1,921 (–43-4,024) first episodes of LRTI and pneumonia, respectively. Estimates were similar when considering all new-onset LRTI and pneumonia cases rather than first episodes only.

**Table 4:** Burden of medically-attended lower respiratory tract infection and pneumonia prevented by PCV13.

| Endpoint | Observed rate among<br>the vaccinated | Counterfactual rate,<br>without PCV13 protection<br>(95% CI) | Averted<br>incidence (95%<br>CI) | Number needed to vaccinate (95%<br>CI) |  | Cases averted<br>among older<br>adults in KPSC<br>cohort, 2018-19<br>season (95% CI)<br>N=460,750 |
| --- | --- | --- | --- | --- | --- | --- |
|  | Per 100 person-years | Per 100 person-years | Per 100 person-years | Preventing 1<br>case over 1 year | Preventing 1 case<br>over 5 years |  |
| Medically-attended<br>LRTI |  |  |  |  |  |  |
| First events only | 7.0 | 7.7 (7.1, 8.4) | 0.7 (0.2, 1.4) | 134 (71, 580) | 27 (14, 116) | 2822 (552, 5195) |
| Any LRTI<sup>1</sup> | 7.2 | 8.0 (7.4, 8.6) | 0.8 (0.2, 1.4) | 131 (69, 564) | 26 (14, 113) | 2903 (568, 5345) |
| Pneumonia |  |  |  |  |  |  |
| First events only | 4.9 | 5.4 (4.9, 5.9) | 0.5 (0.0, 1.0) | 210 (–484, 1341) | 42 (–97, 268) | 1921 (–43, 4024) |
| Any pneumonia<sup>1</sup> | 5.2 | 5.7 (5.2, 6.3) | 0.5 (0.0, 1.1) | 198 (–455, 1262) | 40 (–91, 252) | 2042 (–46, 4277) |

1. Analyses for any LRTI and any pneumonia include all diagnoses not preceded by a prior LRTI or pneumonia diagnosis within 30 days.

## DISCUSSION

Adults aged ≥65 years enrolled in KPSC health plans experienced 9.9% and 8.8% lower incidence of LRTI and pneumonia, respectively, in the years following PCV13 receipt as compared to the years before they were vaccinated. In contrast, we did not observe lower incidence of LRTI and pneumonia following PPSV23 receipt. Analyses considering other adult vaccines as positive and negative control exposures supported the validity of our findings; seasonal influenza vaccination was protective against LRTI and pneumonia, whereas zoster vaccination was not associated with protection against either endpoint. Over a five-year time horizon, ≥1 episode of LRTI and pneumonia, respectively, was prevented for every 27 and 42 adults aged ≥65 years who received PCV13. These findings suggest that under current levels of circulation of vaccine-targeted pneumococcal serotypes, and in the context of long-standing immunization with PPSV23, receipt of PCV13 remains beneficial for older adults.

Our findings indicate that despite widespread and longstanding pediatric PCV13 use in the United States, PCV13-preventable LRTI and pneumonia remain common among older adults. This result is consistent with surveillance data indicating that 49% of US adult pneumococcal pneumonia cases are attributable to PCV13 serotypes.^2^ Serotypes 3 and 19A remain the most prominent contributors to this burden, accounting for 12% and 14% of pneumococcal pneumonia cases, respectively. Previous analyses have reported weaker effectiveness of PCV13 against carriage of these serotypes among children,^16^ potentially accounting for their persisting high prevalence.^17^ In alignment with our findings, these data suggest that an unmet medical need for direct protection against vaccine-serotype pneumonia persists among adults in the context of pediatric immunization. Studies with serotype-specific non-bacteremic pneumonia endpoints have estimated 53% and 59% direct protection against disease due to serotypes 3 and 19A, respectively, among older adults,^18,19^ confirming the value of direct immunization to prevent disease due to these serotypes as they continue to circulate in the community.

While data on the impact of adult PCV13 use against respiratory infection outcomes are limited, estimates from our study are similar to several others. In the Netherlands, PCV13 efficacy against clinically-diagnosed community-acquired pneumonia was estimated at 8% and 10% within strata of older adults with and without known pneumococcal disease risk factors.^20^ Similarly, a cohort study in Germany reported 12% VE against all-cause inpatient and outpatient pneumonia among adults aged ≥60 years, with PCV13 use preventing 0.2 pneumonia cases per 100 persons annually in this population (with a background incidence of 1.6 pneumonia cases per 100 vaccinated persons annually, in contrast to 4.0 episodes per 100 vaccinated persons annually in our study).^21^ Last, in an analysis of US Medicare beneficiaries aged ≥65 years, estimates of PCV13 direct effects against community-acquired pneumonia spanned 6-11%.^22^

These reported reductions in pneumonia incidence among PCV13 recipients suggest the proportion of cases preventable by PCV13 may be under-estimated by available urinary antigen detection (UAD) assays frequently used for serotyping in epidemiologic studies.^23,24^ For example, a recent US study attributed 4.2% of all-cause pneumonia cases among older adults to PCV13-serotype pneumococci based on UAD assay findings,^2^ whereas our study and others^20–22^ estimate that PCV13 prevents 6-12% of cases. Several hypotheses may account for this gap. First, because UAD assays are validated against blood culture-confirmed bacteremic pneumonia as a gold standard, assay sensitivity may be lower in non-bacteremic pneumonia cases potentially associated with lower urinary antigen levels.^25^ Second, synergistic interactions of pneumococci with other microbiota in the upper respiratory tract may result in viruses or other pathogens—rather than pneumococci—ultimately progressing to the lung.^26,27^ In either scenario, performance characteristics of UAD assays remain unknown for pneumonia cases where pneumococci play a role in the causal pathway without causing bloodstream infection.

Limitations of our study should be considered. While observational studies face risk of bias, comprehensive data capture within the KPSC healthcare system provided an opportunity to assess effects of other adult vaccinations as a validation strategy.^11^ Use of a self-matched inference framework mitigated bias driven by differences in risk and healthcare seeking among PCV13 recipients and non-recipients.^28^ However, this strategy constrained the analysis population to 42,700 individuals, limiting statistical power. While recommendations for older adults to receive PCV13 were implemented in 2015,^29^ a change from ICD-9-CM to ICD-10-CM coding during the 2015-16 respiratory season precluded inclusion of earlier data.^10^ Use of stringently-defined clinical endpoints would facilitate comparison of findings across studies,^30^ but is infeasible in real-world effectiveness studies like ours when addressing prevalent outcomes such as LRTI and pneumonia, which are diagnosed across outpatient, emergency department, and inpatient settings.

Our study identifies reduced incidence of all-cause LRTI and pneumonia following PCV13 receipt among US adults aged ≥65 years. These results suggest a need for direct, vaccine-conferred protection among older adults despite the substantial impact of pediatric PCV13 programs on vaccine-serotype pneumococcal circulation and disease across all age groups. Updated assessments of the health economic value of adult PCV13 use are warranted in light of findings from our study and others providing real-world evidence of vaccine direct effects against all-cause, primarily non-invasive disease endpoints.

## Data Availability

Data are not available for public sharing.

## ACNOWLEDGMENTS

The authors thank Magdalena Pomichowski and Ronika Alexander Parrish for support in this project. The study was funded by Pfizer, Inc.

---

**Table S1:** ICD-10-CM codes used for outcome definitions.

| ICD-10-CM code | Definition | Outcome |  |
| --- | --- | --- | --- |
|  |  | LRTI | Pneumonia |
| J09 | Influenza due to certain identified influenza viruses |  |  |
| J09.X1 | With pneumonia | X | X |
| J09.X2 | With other respiratory manifestations | X |  |
| J09.X3 | With gastrointestinal manifestations | X |  |
| J09.X9 | With other manifestations | X |  |
| J10 | Influenza due to other identified influenza virus |  |  |
| J10.00 | With unspecified type of pneumonia | X | X |
| J10.01 | With the same other identified influenza virus pneumonia | X | X |
| J10.08 | With other unspecified pneumonia | X | X |
| J10.1 | With other respiratory manifestations | X |  |
| J10.2 | With gastrointestinal manifestations | X |  |
| J10.81 | With encephalopathy | X |  |
| J10.82 | With myocarditis | X |  |
| J10.83 | With otitis media | X |  |
| J10.89 | With other manifestations | X |  |
| J11 | Influenza due to unidentified influenza virus |  |  |
| J11.00 | With unspecified type of pneumonia | X | X |
| J11.08 | With specified pneumonia | X | X |
| J11.1 | With other respiratory manifestations | X |  |
| J11.2 | With gastrointestinal manifestations | X |  |
| J11.81 | With encephalopathy | X |  |
| J11.82 | With myocarditis | X |  |
| J11.83 | With otitis media | X |  |
| J11.89 | With other manifestations | X |  |
| J12– | Viral pneumonia, not elsewhere classified | X | X |
| J13– | Pneumonia due to *Streptococcus pneumoniae* | X | X |
| J14 | Pneumonia due to *Haemophilus influenzae* | X | X |
| J15– | Bacterial pneumonia, not elsewhere classified | X | X |
| J16– | Pneumonia due to other infectious organisms, not elsewhere classified | X | X |
| J17 | Pneumonia in diseases classified elsewhere | X | X |
| J18– (excl. J18.2) | Pneumonia, unspecified organism (excl. hypostatic pneumonia, unspecified organism) | X | X |
| J20– | Acute bronchitis | X |  |
| J21– | Acute bronchiolitis | X |  |
| J22 | Unspecified acute lower respiratory infection | X |  |

Items marked JX– are nonspecific (non-billable) codes for which the LRTI and pneumonia classifications indicated in the right columns encompass all sub-codes.

**Table S2:** ICD-10-CM codes used to assign comorbid conditions.

| Medical condition | ICD-10-CM codes |
| --- | --- |
| Asthma | J45.x, R06.x |
| Congestive heart failure | I09.9, I11.0, I13.0, I13.2, I25.5, I42.0, I42.5-I42.9, I43.x, I50.x, P29.0 |
| Peripheral vascular disease | I70.x, I71.x, I73.1, I73.8, I73.9, I77.1, I79.0, I79.2, K55.1, K55.8, K55.9, Z95.8, Z95.9 |
| Cerebrovascular disease | G45.x, G46.x, H34.0, I60.x-I69.x |
| COPD | I27.8, I27.9, J40.x-J47.x, J60.x-J67.x, J68.4, J70.1, J70.3 |
| Diabetes | E10.0, E10.1, E10.6, E10.8, E10.9, E11.0, E11.1, E11.6, E11.8, E11.9, E12.0, E12.1, E12.6, E12.8, E12.9, E13.0, E13.1, E13.6, E13.8, E13.9, E14.0, E14.1, E14.6, E14.8, E14.9, E10.2-E10.5, E10.7, E11.2-E11.5, E11.7, E12.2-E12.5, E12.7, E13.2-E13.5, E13.7, E14.2-E14.5, E14.7 |
| Moderate/severe liver disease | I85.0, I85.9, I86.4, I98.2, K70.4, K71.1, K72.1, K72.9, K76.5, K76.6, K76.7 |
| Renal disease | I12.0, I13.1, N03.2-N03.7, N05.2-N05.7, N18.x, N19.x, N25.0, Z49.0-Z49.2, Z94.0, Z99.2 |
| Malignancy | C00.x-C26.x, C30.x-C34.x, C37.x-C41.x, C43.x, C45.x-C58.x, C60.x-C76.x, C81.x-C85.x, C88.x, C90.x-C97.x |
| Metastatic tumor | C77.x-C80.x |
| Other immune conditions | D47.4, D57.x, D68.x, D70.x, D71, D72.x-D76.x, D77, D80.x-D84.x, D89.x, I88.x, M35.9, R75, R76.x |
| Rheumatic/inflammatory disease | M05.x, M06.x, M31.5, M32.x-M35.x, M36.0 |
| AIDS/HIV | B20.x, Z21.x (or identified from HIV Registry) |
| Organ transplant | Identified by National Transplant Services Directory |
| Congestive Heart Failure | I09.9, I11.0, I13.0, I13.2, I25.5, I42.0, I42.5-I42.9, I43.x, I50.x, P29.0 |
| Myocardial Infarction | I21.x, I22.x, I25.2 |

**Table S3:** ACIP high-risk and at-risk status definitions.

| **High-risk** | **At-risk** |
| --- | --- |
| Presence of any of the following conditions:<br>• Congenital or acquired immunodeficiency<br>• Congenital or acquired asplenia<br>• Sickle cell disease/other hemoglobinopathies<br>• HIV infection<br>• Chronic renal failure<br>• Nephrotic syndrome<br>• Leukemia<br>• Lymphoma<br>• Hodgkin's disease<br>• Generalized malignancy<br>• Iatrogenic immunosuppression<br>• Solid organ transplantation<br>• Multiple myeloma<br>• Cerebrospinal fluid leak<br>• Cochlear implant | Presence of any of the following conditions, for individuals without immunosuppressed or immunocompromised status:<br>• Diabetes<br>• Chronic heart disease<br>• Chronic lung disease<br>• Chronic liver disease<br>• Alcoholism<br>• Smoking |

**Table S4:** Attributes of individuals aged ≥65 years within the full Kaiser Permanente cohort who received and those who had not received 13-valent pneumococcal conjugate vaccine as of 1 September, 2016 and 1 September, 2019.

| Attribute |  | Attributes of population by history of PCV13 receipt on 1 September 2016 |  | Attributes of population by history of PCV13 receipt on 1 September 2019 |  |
| --- | --- | --- | --- | --- | --- |
|  |  | PCV13 not received, n (%) | PCV13 received, n (%) | PCV13 not received, n (%) | PCV13 received, n (%) |
|  |  | N=96,938 | N=371,869 | N=73,377 | N=476,873 |
| Age | 65-69y | 42,095 (43.4) | 122,595 (33.0) | 37,726 (51.4) | 143,645 (30.1) |
|  | 70-74y | 24,382 (25.2) | 98,493 (26.5) | 17,879 (24.4) | 139,161 (29.2) |
|  | 75-79y | 14,002 (14.4) | 69,324 (18.6) | 9,030 (12.3) | 89,731 (18.8) |
|  | 80-84y | 8,654 (8.9) | 45,345 (12.2) | 4,913 (6.7) | 57,454 (12.0) |
|  | 85-89y | 4,743 (4.9) | 24,332 (6.5) | 2,456 (3.3) | 31,202 (6.5) |
|  | ≥90y | 3,062 (3.2) | 11,780 (3.2) | 1,373 (1.9) | 15,680 (3.3) |
| Sex | Female | 53,830 (55.5) | 204,906 (55.1) | 40,265 (54.9) | 263,468 (55.2) |
|  | Male | 43,108 (44.5) | 166,963 (44.9) | 33,112 (45.1) | 213,405 (44.8) |
| Race and ethnicity | White, non-Hispanic | 48,054 (49.6) | 195,579 (52.6) | 34,627 (47.2) | 235,719 (49.4) |
|  | Black, non-Hispanic | 12,756 (13.2) | 33,737 (9.1) | 10,169 (13.9) | 42,527 (8.9) |
|  | Asian/Pacific Islander, non-Hispanic | 9,048 (9.3) | 44,443 (12.0) | 6,167 (8.4) | 60,160 (12.6) |
|  | Hispanic | 21,760 (22.4) | 92,892 (25.0) | 16,176 (22.0) | 128,942 (27.0) |
|  | Native American/Alaska Native | 271 (0.3) | 878 (0.2) | 217 (0.3) | 1,162 (0.2) |
|  | Mixed race | 141 (0.1) | 489 (0.1) | 118 (0.2) | 679 (0.1) |
|  | Other/unknown | 4,908 (5.1) | 3,851 (1.0) | 5,903 (8.0) | 7,684 (1.6) |
| Medical history | Myocardial infarction | 4,138 (4.3) | 23,892 (6.4) | 2,136 (2.9) | 25,388 (5.3) |
|  | Hypertension | 20,953 (21.6) | 138,446 (37.2) | 29,280 (39.9) | 317,105 (66.5) |
|  | Congestive heart failure | 3,573 (3.7) | 21,444 (5.8) | 2,129 (2.9) | 32,155 (6.7) |
|  | Peripheral vascular disease | 21,776 (22.5) | 145,951 (39.2) | 12,068 (16.4) | 191,168 (40.1) |
|  | Cerebrovascular disease | 4,354 (4.5) | 22,934 (6.2) | 2,319 (3.2) | 29,841 (6.3) |
|  | Asthma | 1,678 (1.7) | 13,883 (3.7) | 4,185 (5.7) | 53,713 (11.3) |
|  | Chronic pulmonary disease | 10,269 (10.6) | 70,477 (19.0) | 6,717 (9.2) | 90,509 (19.0) |
|  | Uncomplicated diabetes | 9,723 (10.0) | 50,321 (13.5) | 8,073 (11.0) | 65,844 (13.8) |
|  | Complicated diabetes | 9,851 (10.2) | 65,705 (17.7) | 6,456 (8.8) | 89,0529 (18.7) |
|  | Renal disease | 13,166 (13.6) | 86,618 (23.3) | 6,500 (8.9) | 96,630 (20.3) |
|  | Moderate to severe liver disease | 196 (0.2) | 1,178 (0.3) | 143 (0.2) | 1,802 (0.4) |
|  | Malignancy | 3,617 (3.7) | 19,435 (5.2) | 2,262 (3.1) | 26,105 (5.5) |
|  | Tumor | 1,054 (1.1) | 4,878 (1.3) | 668 (0.9) | 7,370 (1.5) |
|  | Rheumatic or connective tissue disease | 1,767 (1.8) | 11,634 (3.1) | 1,053 (1.4) | 12,798 (2.7) |
|  | Hypothyroid | 4,030 (4.2) | 25,737 (6.9) | 6,095 (8.3) | 70,520 (14.8) |
|  | HIV/AIDS | 13 (<0.1) | 38 (<0.1) | 9 (<0.1) | 98 (<0.1) |
|  | Organ transplant | 82 (0.1) | 385 (0.1) | 51 (0.1) | 589 (0.1) |
|  | Other immune deficiency or suppression | 754 (0.8) | 5,018 (1.3) | 1,810 (2.5) | 22,042 (4.6) |
|  | Depression | 3,578 (3.7) | 25,150 (6.8) | 5,243 (7.1) | 75,239 (15.8) |
| Body mass index | Underweight | 1,099 (1.1) | 3,794 (1.0) | 701 (1.0) | 4,593 (1.0) |
|  | Normal weight | 16,382 (16.9) | 72,129 (19.4) | 11,155 (15.2) | 87,602 (18.4) |
|  | Overweight | 19,735 (20.4) | 96,047 (25.8) | 13,811 (18.8) | 118,381 (24.8) |
|  | Obese | 10,526 (10.9) | 52,354 (14.1) | 7,568 (10.3) | 66,187 (13.9) |
|  | Morbidly obese | 5,339 (5.5) | 28,317 (7.6) | 4,287 (5.8) | 37,087 (7.8) |
|  | Unknown | 43,857 (45.2) | 119,228 (32.1) | 35,855 (48.9) | 163,023 (34.2) |
| Cigarette smoking | Current smoker | 5,075 (5.2) | 15,669 (4.2) | 3,510 (4.8) | 17,655 (3.7) |
|  | Former smoker | 25,578 (26.4) | 141,623 (38.1) | 15,443 (21.0) | 166,511 (34.9) |
|  | Never smoked | 46,814 (48.3) | 206,314 (55.5) | 31,272 (42.6) | 268,111 (56.2) |
|  | Unknown | 19,471 (20.1) | 8,263 (2.2) | 23,152 (31.6) | 24,596 (5.2) |
| ACIP pneumococcal |  |  |  |  |  |
| disease risk designation |  |  |  |  |  |
|  | Normal risk | 60,860 (62.8) | 168,563 (45.3) | 40,655 (55.4) | 155,579 (32.6) |
|  | At risk | 29,802 (30.7) | 163,259 (43.9) | 26,385 (36.0) | 238,219 (50.0) |
|  | High risk | 6,276 (6.5) | 40,047 (10.8) | 6,337 (8.6) | 83,075 (17.4) |
| Other vaccines |  |  |  |  |  |
|  | PPSV23<sup>1</sup> | 70,022 (72.2) | 358,143 (96.3) | 30,311 (41.3) | 432,469 (90.7) |
|  | Prior year seasonal influenza vaccination<sup>2</sup> | 23,020/80,152 (28.7) | 200,712/329,072 (61.0) | 9,295/55,765 (16.7) | 357,931/434,711 (82.3) |
|  | Any zoster vaccine<sup>1</sup> | 20,574 (21.2) | 182,346 (49.0) | 9,268 (12.6) | 226,633 (47.5) |
| Prior-year healthcare utilization |  |  |  |  |  |
|  | Outpatient visits, mean (±SD) | 14.5 (15.3) | 22.5 (18.1) | 13.4 (15.5) | 23.2 (19.6) |
|  | Emergency department visits, mean (±SD) | 0.3 (0.8) | 0.4 (0.9) | 0.2 (0.7) | 0.4 (1.0) |
|  | Inpatient admissions, mean (±SD) | 0.09 (0.4) | 0.13 (0.5) | 0.05 (0.3) | 0.11 (0.4) |
| Socioeconomic status |  |  |  |  |  |
| | Census tract median household income, mean (±SD), in \$1000s | 70.5 (29.0) | 72.6 (29.4) | 81.5 (34.0) | 83.9 (34.7) |

Counts include all KPSC members aged ≥65 years with at least one year of enrollment prior to the start date of each season. Individuals who received ACIP-discordant pneumococcal vaccine sequences are excluded.
1. Indicates the number of individuals who ever received PPSV23 and zoster vaccine previously. 2. Proportions calculated among total population enrolled during the prior year.

**Table S5:**
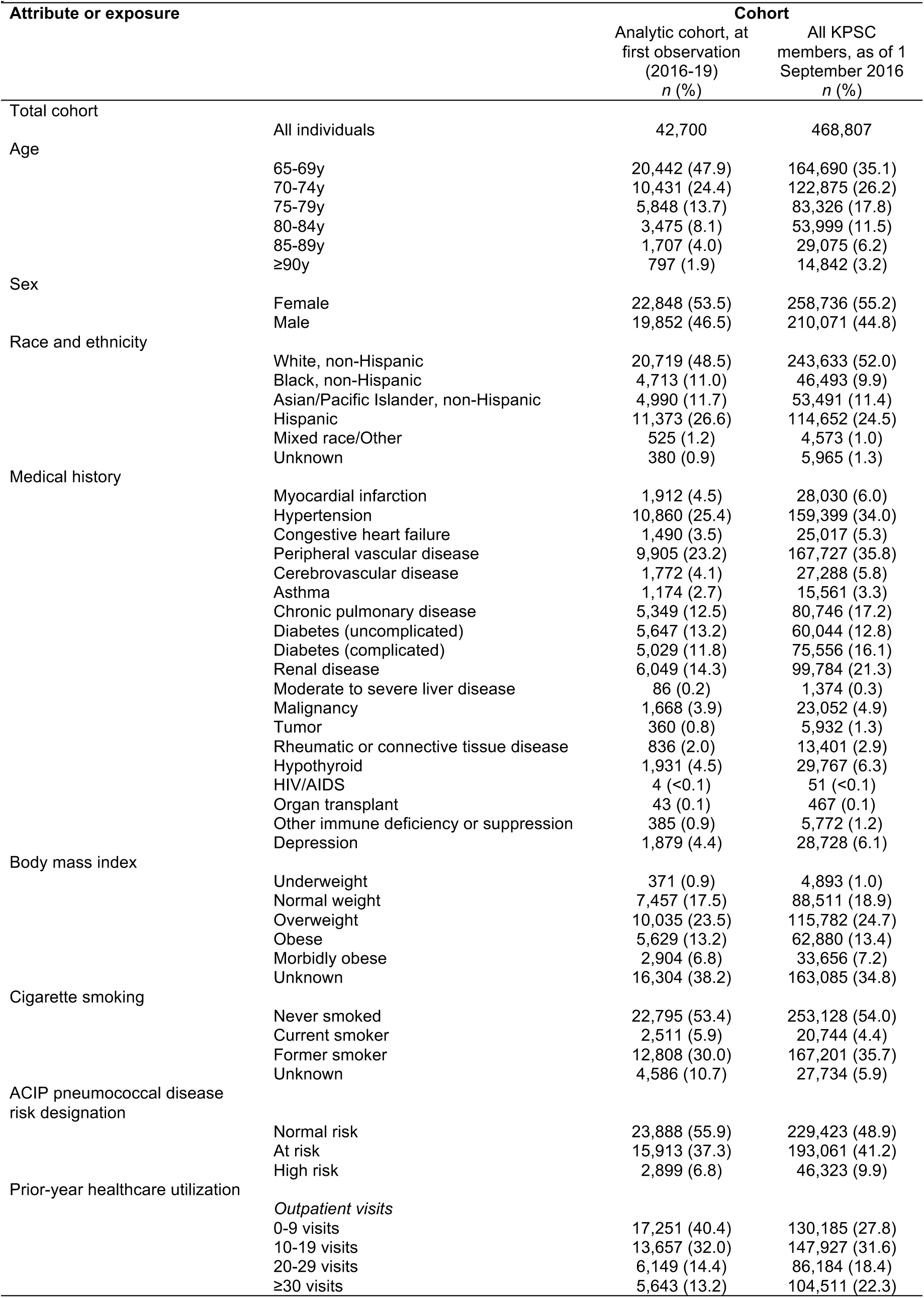
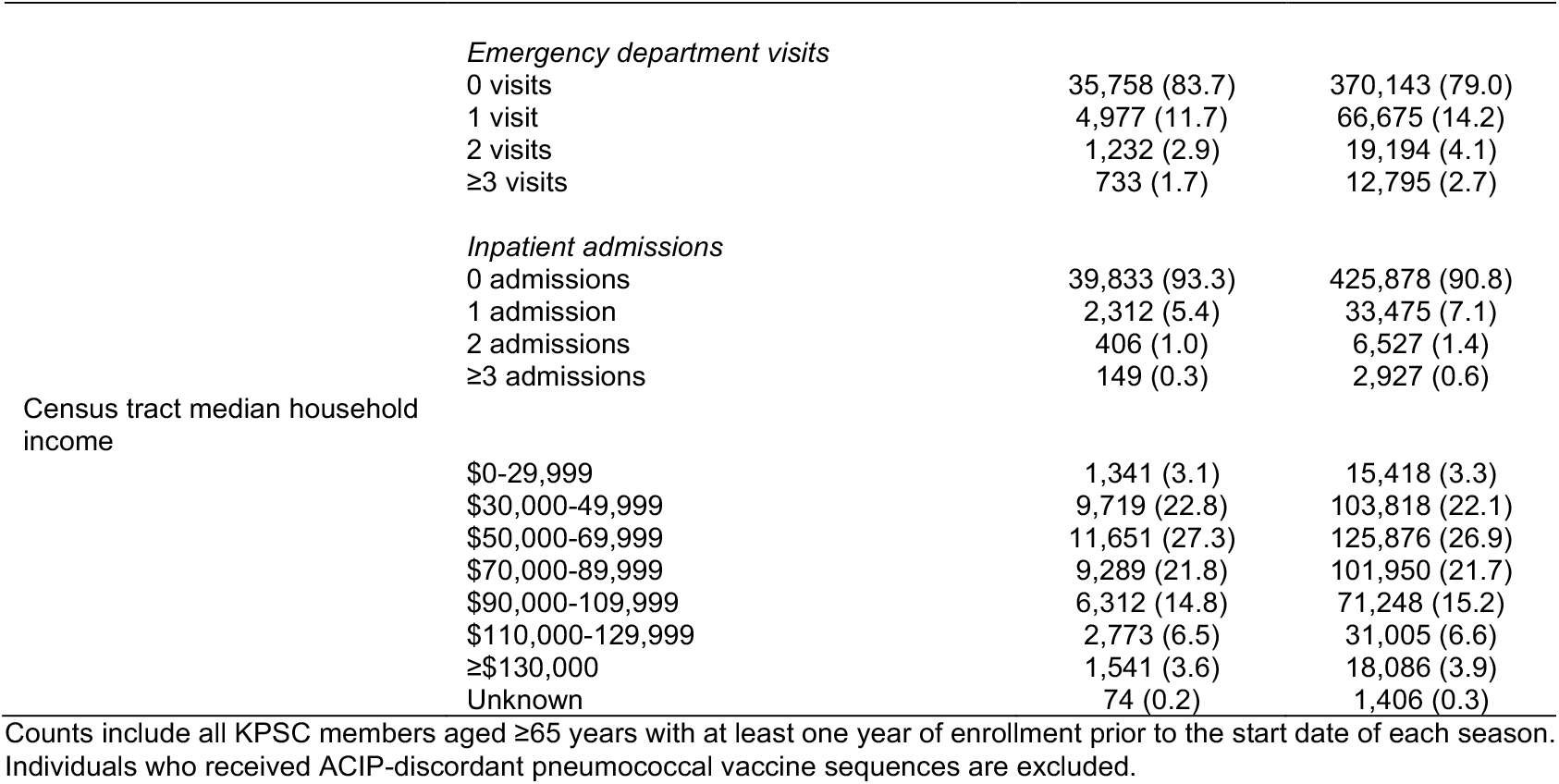
Comparison of members of the analytic cohort to KPSC cohort members aged ≥65 years.

| Attribute or exposure |  | Cohort |  |
| --- | --- | --- | --- |
|  |  | Analytic cohort, at first observation (2016-19)<br>*n* (%) | All KPSC members, as of 1 September 2016<br>*n* (%) |
| Total cohort | All individuals | 42,700 | 468,807 |
| Age |  |  |  |
|  | 65-69y | 20,442 (47.9) | 164,690 (35.1) |
|  | 70-74y | 10,431 (24.4) | 122,875 (26.2) |
|  | 75-79y | 5,848 (13.7) | 83,326 (17.8) |
|  | 80-84y | 3,475 (8.1) | 53,999 (11.5) |
|  | 85-89y | 1,707 (4.0) | 29,075 (6.2) |
|  | ≥90y | 797 (1.9) | 14,842 (3.2) |
| Sex |  |  |  |
|  | Female | 22,848 (53.5) | 258,736 (55.2) |
|  | Male | 19,852 (46.5) | 210,071 (44.8) |
| Race and ethnicity |  |  |  |
|  | White, non-Hispanic | 20,719 (48.5) | 243,633 (52.0) |
|  | Black, non-Hispanic | 4,713 (11.0) | 46,493 (9.9) |
|  | Asian/Pacific Islander, non-Hispanic | 4,990 (11.7) | 53,491 (11.4) |
|  | Hispanic | 11,373 (26.6) | 114,652 (24.5) |
|  | Mixed race/Other | 525 (1.2) | 4,573 (1.0) |
|  | Unknown | 380 (0.9) | 5,965 (1.3) |
| Medical history |  |  |  |
|  | Myocardial infarction | 1,912 (4.5) | 28,030 (6.0) |
|  | Hypertension | 10,860 (25.4) | 159,399 (34.0) |
|  | Congestive heart failure | 1,490 (3.5) | 25,017 (5.3) |
|  | Peripheral vascular disease | 9,905 (23.2) | 167,727 (35.8) |
|  | Cerebrovascular disease | 1,772 (4.1) | 27,288 (5.8) |
|  | Asthma | 1,174 (2.7) | 15,561 (3.3) |
|  | Chronic pulmonary disease | 5,349 (12.5) | 80,746 (17.2) |
|  | Diabetes (uncomplicated) | 5,647 (13.2) | 60,044 (12.8) |
|  | Diabetes (complicated) | 5,029 (11.8) | 75,556 (16.1) |
|  | Renal disease | 6,049 (14.3) | 99,784 (21.3) |
|  | Moderate to severe liver disease | 86 (0.2) | 1,374 (0.3) |
|  | Malignancy | 1,668 (3.9) | 23,052 (4.9) |
|  | Tumor | 360 (0.8) | 5,932 (1.3) |
|  | Rheumatic or connective tissue disease | 836 (2.0) | 13,401 (2.9) |
|  | Hypothyroid | 1,931 (4.5) | 29,767 (6.3) |
|  | HIV/AIDS | 4 (<0.1) | 51 (<0.1) |
|  | Organ transplant | 43 (0.1) | 467 (0.1) |
|  | Other immune deficiency or suppression | 385 (0.9) | 5,772 (1.2) |
|  | Depression | 1,879 (4.4) | 28,728 (6.1) |
| Body mass index |  |  |  |
|  | Underweight | 371 (0.9) | 4,893 (1.0) |
|  | Normal weight | 7,457 (17.5) | 88,511 (18.9) |
|  | Overweight | 10,035 (23.5) | 115,782 (24.7) |
|  | Obese | 5,629 (13.2) | 62,880 (13.4) |
|  | Morbidly obese | 2,904 (6.8) | 33,656 (7.2) |
|  | Unknown | 16,304 (38.2) | 163,085 (34.8) |
| Cigarette smoking |  |  |  |
|  | Never smoked | 22,795 (53.4) | 253,128 (54.0) |
|  | Current smoker | 2,511 (5.9) | 20,744 (4.4) |
|  | Former smoker | 12,808 (30.0) | 167,201 (35.7) |
|  | Unknown | 4,586 (10.7) | 27,734 (5.9) |
| ACIP pneumococcal disease risk designation |  |  |  |
|  | Normal risk | 23,888 (55.9) | 229,423 (48.9) |
|  | At risk | 15,913 (37.3) | 193,061 (41.2) |
|  | High risk | 2,899 (6.8) | 46,323 (9.9) |
| Prior-year healthcare utilization |  |  |  |
|  | *Outpatient visits* |  |  |
|  | 0-9 visits | 17,251 (40.4) | 130,185 (27.8) |
|  | 10-19 visits | 13,657 (32.0) | 147,927 (31.6) |
|  | 20-29 visits | 6,149 (14.4) | 86,184 (18.4) |
|  | ≥30 visits | 5,643 (13.2) | 104,511 (22.3) |
|  | *Emergency department visits* |  |  |
|  | 0 visits | 35,758 (83.7) | 370,143 (79.0) |
|  | 1 visit | 4,977 (11.7) | 66,675 (14.2) |
|  | 2 visits | 1,232 (2.9) | 19,194 (4.1) |
|  | ≥3 visits | 733 (1.7) | 12,795 (2.7) |
|  | *Inpatient admissions* |  |  |
|  | 0 admissions | 39,833 (93.3) | 425,878 (90.8) |
|  | 1 admission | 2,312 (5.4) | 33,475 (7.1) |
|  | 2 admissions | 406 (1.0) | 6,527 (1.4) |
|  | ≥3 admissions | 149 (0.3) | 2,927 (0.6) |
| Census tract median household income |  |  |  |
| | \$0-29,999 | 1,341 (3.1) | 15,418 (3.3) |
| | \$30,000-49,999 | 9,719 (22.8) | 103,818 (22.1) |
| | \$50,000-69,999 | 11,651 (27.3) | 125,876 (26.9) |
| | \$70,000-89,999 | 9,289 (21.8) | 101,950 (21.7) |
| | \$90,000-109,999 | 6,312 (14.8) | 71,248 (15.2) |
| | \$110,000-129,999 | 2,773 (6.5) | 31,005 (6.6) |
| | ≥\$130,000 | 1,541 (3.6) | 18,086 (3.9) |
|  | Unknown | 74 (0.2) | 1,406 (0.3) |

Counts include all KPSC members aged ≥65 years with at least one year of enrollment prior to the start date of each season. Individuals who received ACIP-discordant pneumococcal vaccine sequences are excluded.

**Table S6:** Attributes of individuals aged ≥65 years within the full KSPC cohort who received and those who had not received 23-valent pneumococcal polysaccharide vaccine as of 1 September 2016 and 1 September 2019.

| Attribute |  | Attributes of population by history of PPSV23 receipt on 1 September 2016 |  | Attributes of population by history of PPSV23 receipt on 1 September 2019 |  |
| --- | --- | --- | --- | --- | --- |
|  |  | PPSV23 not received, *n* (%)<br>*N*=40,642 | PPSV23 received, *n* (%)<br>*N*=428,165 | PPSV23 not received, *n* (%)<br>*N*=87,470 | PPSV23 received, *n* (%)<br>*N*=462,780 |
| Age | 65-69y | 25,591 (63.0) | 139,099 (32.5) | 57,465 (65.7) | 123,906 (26.8) |
|  | 70-74y | 7,615 (18.7) | 115,260 (26.9) | 17,415 (19.9) | 139,625 (30.2) |
|  | 75-79y | 3,320 (7.9) | 80,096 (18.7) | 6,554 (7.5) | 92,207 (19.9) |
|  | 80-84y | 2,268 (5.6) | 51,731 (12.1) | 3,135 (3.6) | 59,232 (12.8) |
|  | 85-89y | 1,242 (3.1) | 27,833 (6.5) | 1,868 (2.1) | 31,790 (6.9) |
|  | ≥90y | 696 (1.7) | 14,146 (3.3) | 1,033 (1.2) | 16,020 (3.5) |
| Sex | Female | 23,378 (57.5) | 235,538 (55.0) | 48,138 (55.0) | 255,595 (55.2) |
|  | Male | 17,264 (42.5) | 192,807 (45.0) | 39,332 (45.0) | 207,185 (44.8) |
| Race and ethnicity | White, non-Hispanic | 20,580 (50.6) | 223,053 (52.1) | 41,298 (47.2) | 229,048 (49.5) |
|  | Black, non-Hispanic | 5,265 (13.0) | 41,228 (9.6) | 9,595 (11.0) | 43,101 (9.3) |
|  | Asian/Pacific Islander, non-Hispanic | 3,387 (8.3) | 50,104 (11.7) | 8,679 (9.9) | 57,648 (12.5) |
|  | Hispanic | 7,534 (18.5) | 107,118 (25.0) | 20,879 (23.9) | 124,239 (26.8) |
|  | Native American/Alaska Native | 109 (0.3) | 1,040 (0.2) | 236 (0.3) | 1,143 (0.2) |
|  | Mixed race | 66 (0.2) | 564 (0.1) | 146 (0.2) | 651 (0.1) |
|  | Other/unknown | 3,701 (9.1) | 5,058 (1.2) | 6,637 (7.6) | 6,950 (1.5) |
| Medical history | Myocardial infarction | 959 (2.4) | 2,701 (6.3) | 1,653 (1.9) | 25,871 (5.6) |
|  | Hypertension | 7,698 (18.9) | 151,701 (35.4) | 33,729 (38.6) | 312,656 (67.6) |
|  | Congestive heart failure | 730 (1.8) | 24,287 (5.7) | 1,806 (2.1) | 32,478 (7.0) |
|  | Peripheral vascular disease | 5,523 (13.6) | 162,204 (37.9) | 11,638 (13.3) | 191,598 (41.4) |
|  | Cerebrovascular disease | 1,084 (2.7) | 26,204 (6.1) | 2,172 (2.5) | 29,988 (6.5) |
|  | Asthma | 550 (1.4) | 15,011 (3.5) | 3,937 (4.5) | 53,961 (11.7) |
|  | Chronic pulmonary disease | 2,807 (6.9) | 77,939 (18.2) | 6,345 (7.3) | 90,881 (19.6) |
|  | Uncomplicated diabetes | 2,713 (6.7) | 57,331 (13.4) | 6,512 (7.4) | 67,405 (14.6) |
|  | Complicated diabetes | 1,774 (4.4) | 73,782 (17.2) | 3,931 (4.5) | 91,577 (19.8) |
|  | Renal disease | 3,258 (8.0) | 96,526 (22.5) | 5,462 (6.2) | 97,668 (21.1) |
|  | Moderate to severe liver disease | 44 (0.1) | 1,330 (0.3) | 125 (0.1) | 1,820 (0.4) |
|  | Malignancy | 1,276 (3.1) | 21,776 (5.1) | 2,767 (3.2) | 25,600 (5.5) |
|  | Tumor | 338 (0.8) | 5,594 (1.3) | 786 (0.9) | 7,252 (1.6) |
|  | Rheumatic or connective tissue disease | 560 (1.4) | 12,841 (3.0) | 1,077 (1.2) | 12,774 (2.8) |
|  | Hypothyroid | 1,621 (4.0) | 28,146 (6.6) | 7,540 (8.6) | 69,075 (14.9) |
|  | HIV/AIDS | 4 (<0.1) | 47 (<0.1) | 3 (<0.1) | 104 (<0.1) |
|  | Organ transplant | 19 (<0.1) | 448 (0.1) | 31 (<0.1) | 609 (0.1) |
|  | Other immune deficiency or suppression | 287 (0.7) | 5,485 (1.3) | 2,236 (2.6) | 21,616 (4.7) |
|  | Depression | 1,290 (3.2) | 27,438 (6.4) | 6,198 (7.1) | 74,284 (16.1) |
| Body mass index | Underweight | 394 (1.0) | 4,499 (1.1) | 768 (0.9) | 4,526 (1.0) |
|  | Normal weight | 6,866 (16.9) | 81,645 (19.1) | 14,723 (16.8) | 84,034 (18.2) |
|  | Overweight | 8,232 (20.3) | 107,550 (25.1) | 18,930 (21.6) | 113,262 (24.5) |
|  | Obese | 4,252 (10.5) | 58,628 (13.7) | 10,020 (11.5) | 63,735 (13.8) |
|  | Morbidly obese | 2,213 (5.4) | 31,443 (7.3) | 5,194 (5.9) | 36,180 (7.8) |
|  | Unknown | 18,685 (46.0) | 144,400 (33.7) | 37,835 (43.3) | 161,043 (34.8) |
| Cigarette smoking | Current smoker | 1,642 (4.0) | 19,102 (4.5) | 2,885 (3.3) | 18,280 (4.0) |
|  | Former smoker | 9,014 (22.2) | 158,187 (36.9) | 18,220 (20.8) | 163,734 (35.4) |
|  | Never smoked | 20,348 (50.1) | 232,780 (54.4) | 45,331 (51.8) | 254,052 (54.9) |
|  | Unknown | 9,638 (23.7) | 18,096 (4.2) | 21,034 (24.0) | 26,714 (5.8) |
| ACIP pneumococcal |  |  |  |  |  |
| disease risk designation |  |  |  |  |  |
|  | Normal risk | 30,291 (74.5) | 199,132 (46.5) | 55,674 (63.6) | 140,560 (30.4) |
|  | At risk | 8,927 (22.0) | 184,134 (43.0) | 27,441 (31.4) | 237,163 (51.2) |
|  | High risk | 1,424 (3.5) | 44,899 (10.5) | 4,355 (5.0) | 85,057 (18.4) |
| Other vaccines |  |  |  |  |  |
|  | PCV23 | 13,726 (33.8) | 358,143 (83.6) | 44,404 (50.8) | 432,469 (93.5) |
|  | Prior year seasonal influenza vaccination<sup>1</sup> | 829/20,852 (4.0) | 222,697/388,372 (57.3) | 12,808/51,637 (24.8) | 354,019/438,839 (80.7) |
|  | Any zoster vaccine | 6,172 (15.2) | 196,748 (46.0) | 14,689 (16.8) | 221,212 (47.8) |
| Prior-year healthcare utilization |  |  |  |  |  |
|  | Outpatient visits, mean (±SD) | 13.4 (14.5) | 21.5 (17.9) | 14.1 (15.3) | 23.4 (19.8) |
|  | Emergency department visits, mean (±SD) | 0.5 (0.7) | 0.4 (0.9) | 0.2 (0.8) | 0.4 (1.0) |
|  | Inpatient admissions, mean (±SD) | 0.06 (0.3) | 0.13 (0.5) | 0.06 (0.3) | 0.11 (0.4) |
| Socioeconomic status |  |  |  |  |  |
| | Census tract median household income, mean (±SD), in \$1000s | 71.7 (29.8) | 72.2 (29.3) | 83.2 (34.7) | 83.6 (34.6) |

Counts include all KPSC members aged ≥65 years with at least one year of enrollment prior to the start date of each season. Individuals who received ACIP-discordant pneumococcal vaccine sequences are excluded.
1. Proportions calculated among total population enrolled during the prior year.

**Table S7:** Attributes of individuals aged ≥65 years within the full KPSC cohort who received and did not receive zoster and seasonal influenza vaccines.

| Attribute |  | Attributes of population by history of zoster vaccine receipt as of 1 September, 2016 |  | Attributes of population by seasonal influenza vaccine receipt during 2016-17 season |  |
| --- | --- | --- | --- | --- | --- |
|  |  | Zoster vaccine not received, *n* (%) | Zoster vaccine received, *n* (%) | Seasonal influenza vaccine not received, *n* (%) | Seasonal influenza vaccine received, *n* (%) |
|  |  | *N*=265,887 | *N*=202,920 | *N*=149,229 | *N*=319,578 |
| Age | 65-69y | 96,002 (36.1) | 68,688 (33.8) | 60,455 (40.5) | 104,235 (32.6) |
|  | 70-74y | 65,743 (24.7) | 57,132 (28.2) | 38,051 (25.5) | 84,824 (26.5) |
|  | 75-79y | 45,045 (16.9) | 38,281 (18.9) | 22,909 (15.4) | 60,417 (18.9) |
|  | 80-84y | 30,480 (11.5) | 23,519 (11.6) | 14,467 (9.7) | 39,532 (12.4) |
|  | 85-89y | 17,989 (6.8) | 11,086 (5.5) | 8,198 (5.5) | 20,877 (6.5) |
|  | ≥90y | 10,628 (4.0) | 4,214 (2.1) | 5,149 (3.5) | 9,693 (3.0) |
| Sex | Female | 142,462 (53.6) | 116,274 (57.3) | 83,506 (56.0) | 175,230 (54.8) |
|  | Male | 123,425 (46.4) | 86,646 (42.7) | 65,723 (44.0) | 144,348 (45.2) |
| Race and ethnicity | White, non-Hispanic | 121,319 (45.6) | 122,314 (60.3) | 72,060 (48.3) | 171,573 (53.7) |
|  | Black, non-Hispanic | 31,415 (11.8) | 15,078 (7.4) | 19,485 (13.1) | 27,008 (8.5) |
|  | Asian/Pacific Islander, non-Hispanic | 28,180 (10.6) | 25,311 (12.5) | 14,287 (9.6) | 39,204 (12.3) |
|  | Hispanic | 77,344 (29.1) | 37,308 (18.4) | 37,386 (25.1) | 77,266 (24.2) |
|  | Native American/Alaska Native | 715 (0.3) | 434 (0.2) | 406 (0.3) | 743 (0.2) |
|  | Mixed race | 349 (0.1) | 281 (0.1) | 220 (0.1) | 410 (0.1) |
|  | Other/unknown | 6,565 (2.5) | 2,194 (1.1) | 5,385 (3.6) | 3,374 (1.1) |
| Medical history | Myocardial infarction | 16,682 (6.3) | 11,348 (5.6) | 7,660 (5.1) | 20,370 (6.4) |
|  | Hypertension | 89,974 (33.8) | 69,425 (34.2) | 40,528 (27.2) | 118,871 (37.2) |
|  | Congestive heart failure | 15,565 (5.9) | 9,452 (4.7) | 6,850 (4.6) | 18,167 (5.7) |
|  | Peripheral vascular disease | 92,128 (34.6) | 75,599 (37.3) | 46,250 (28.6) | 125,077 (39.1) |
|  | Cerebrovascular disease | 15,976 (6.0) | 11,312 (5.6) | 8,041 (5.4) | 19,247 (6.0) |
|  | Asthma | 8,288 (3.1) | 7,273 (3.6) | 3,483 (2.3) | 12,078 (3.8) |
|  | Chronic pulmonary disease | 44,416 (16.7) | 36,330 (17.9) | 19,999 (13.4) | 60,747 (19.0) |
|  | Uncomplicated diabetes | 35,573 (13.4) | 24,471 (12.1) | 17,457 (11.7) | 42,587 (13.3) |
|  | Complicated diabetes | 45,662 (17.2) | 29,894 (14.7) | 19,186 (12.9) | 56,370 (17.6) |
|  | Renal disease | 56,760 (21.3) | 43,024 (21.2) | 25,275 (16.9) | 74,509 (23.3) |
|  | Moderate to severe liver disease | 927 (0.3) | 447 (0.2) | 396 (0.3) | 978 (0.3) |
|  | Malignancy | 12,968 (4.9) | 10,084 (5.0) | 6,025 (4.0) | 17,027 (5.3) |
|  | Tumor | 3,549 (1.3) | 2,383 (1.2) | 1,724 (1.2) | 4,208 (1.3) |
|  | Rheumatic or connective tissue disease | 8,110 (3.1) | 5,291 (2.6) | 3,174 (2.1) | 10,227 (3.2) |
|  | Hypothyroid | 15,762 (5.9) | 14,005 (6.9) | 7,630 (5.1) | 22,137 (6.9) |
|  | HIV/AIDS | 47 (<0.1) | 4 (<0.1) | 22 (<0.1) | 29 (<0.1) |
|  | Organ transplant | 354 (0.1) | 113 (0.1) | 113 (0.1) | 354 (0.1) |
|  | Other immune deficiency or suppression | 3,196 (1.2) | 2,576 (1.3) | 1,523 (1.0) | 4,249 (1.3) |
|  | Depression | 15,213 (5.7) | 13,515 (6.7) | 7,263 (4.9) | 21,465 (6.7) |
| Body mass index | Underweight | 2,908 (1.1) | 1,985 (1.0) | 1,777 (1.2) | 3,116 (1.0) |
|  | Normal weight | 46,891 (17.6) | 41,620 (20.5) | 26,337 (17.6) | 62,174 (19.5) |
|  | Overweight | 63,452 (23.9) | 52,330 (25.8) | 33,302 (22.3) | 82,480 (25.8) |
|  | Obese | 35,672 (13.4) | 27,208 (13.4) | 18,180 (12.2) | 44,700 (14.0) |
|  | Morbidly obese | 19,529 (7.3) | 14,127 (7.0) | 9,921 (6.6) | 23,735 (7.4) |
|  | Unknown | 97,435 (36.6) | 65,650 (32.4) | 59,712 (40.0) | 103,33 (32.3) |
| Cigarette smoking | Current smoker | 13,786 (5.2) | 6,958 (3.4) | 8,541 (5.7) | 12,203 (3.8) |
|  | Former smoker | 90,174 (33.9) | 77,027 (38.0) | 44,806 (30.0) | 122,395 (38.3) |
|  | Never smoked | 140,105 (52.7) | 113,023 (55.7) | 76,361 (51.2) | 176,767 (55.3) |
|  | Unknown | 21,822 (8.2) | 5,912 (2.9) | 19,521 (13.1) | 8,213 (2.6) |
| ACIP<br>pneumococcal<br>disease risk<br>designation | Normal risk | 127,846 (48.1) | 101,577 (50.1) | 83,761 (56.1) | 145,662 (45.6) |
|  | At risk | 109,894 (41.3) | 83,167 (41.0) | 53,897 (36.1) | 139,164 (43.5) |
|  | High risk | 28,147 (10.6) | 18,176 (9.0) | 11,571 (7.8) | 34,752 (10.9) |
| Other vaccines | PCV13 | 189,523 (71.3) | 182,346 (89.9) | 74,004 (49.6) | 297,865 (93.2) |
|  | PPSV23 | 231,417 (87.0) | 196,748 (97.0) | 117,120 (78.5) | 311,045 (97.3) |
|  | Prior year seasonal influenza<br>vaccination<sup>1</sup> | 105,343/225,323<br>(46.8) | 118,183/183,901<br>(64.3) | 220,150/404,557<br>(54.4) | 194,702/286,211<br>(68.0) |
|  | Any zoster vaccine | -- | -- | 239,895 (26.7) | 163,025 (51.0) |
| Prior-year<br>healthcare<br>utilization | Outpatient visits, mean (±SD) | 19.8 (17.6) | 22.2 (18.0) | 16.8 (16.3) | 22.7 (18.2) |
|  | Emergency department<br>visits, mean (±SD) | 0.4 (0.9) | 0.3 (0.8) | 0.1 (0.5) | 0.3 (0.9) |
|  | Inpatient admissions, mean<br>(±SD) | 0.13 (0.5) | 0.11 (0.4) | 0.11 (0.5) | 0.13 (0.5) |
| Socioeconomic<br>status |  |  |  |  |  |
| | Census tract median<br>household income, mean<br>(±SD), in \$1000s | 69.1 (28.3) | 76.2 (30.2) | 70.1 (28.9) | 73.1 (29.5) |

Counts include all KPSC members aged ≥65 years with at least one year of enrollment prior to the start date of each season. Individuals who received ACIP-discordant pneumococcal vaccine sequences are excluded.
1. Proportions calculated among total population enrolled during the prior year.

**Table S8:**
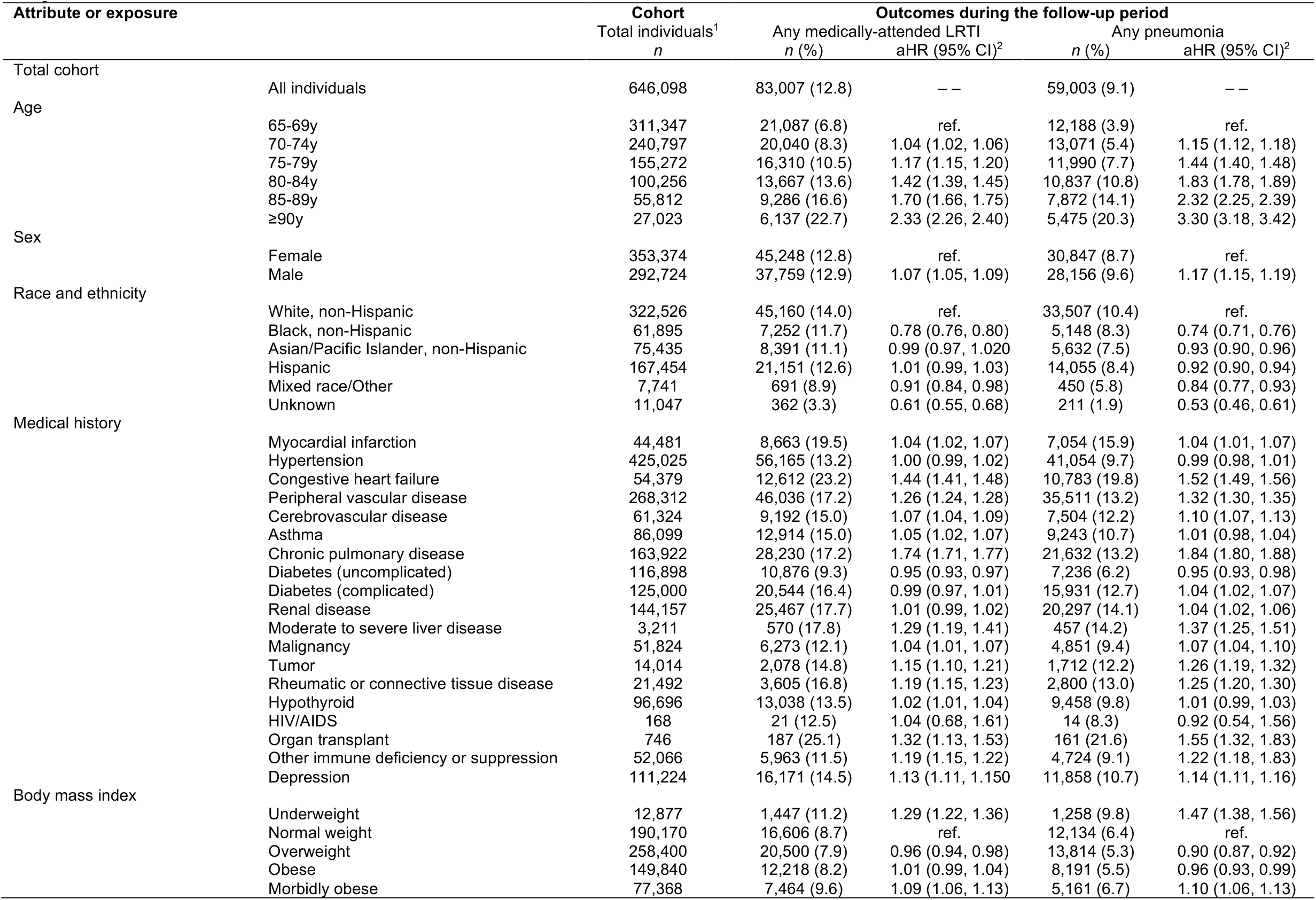
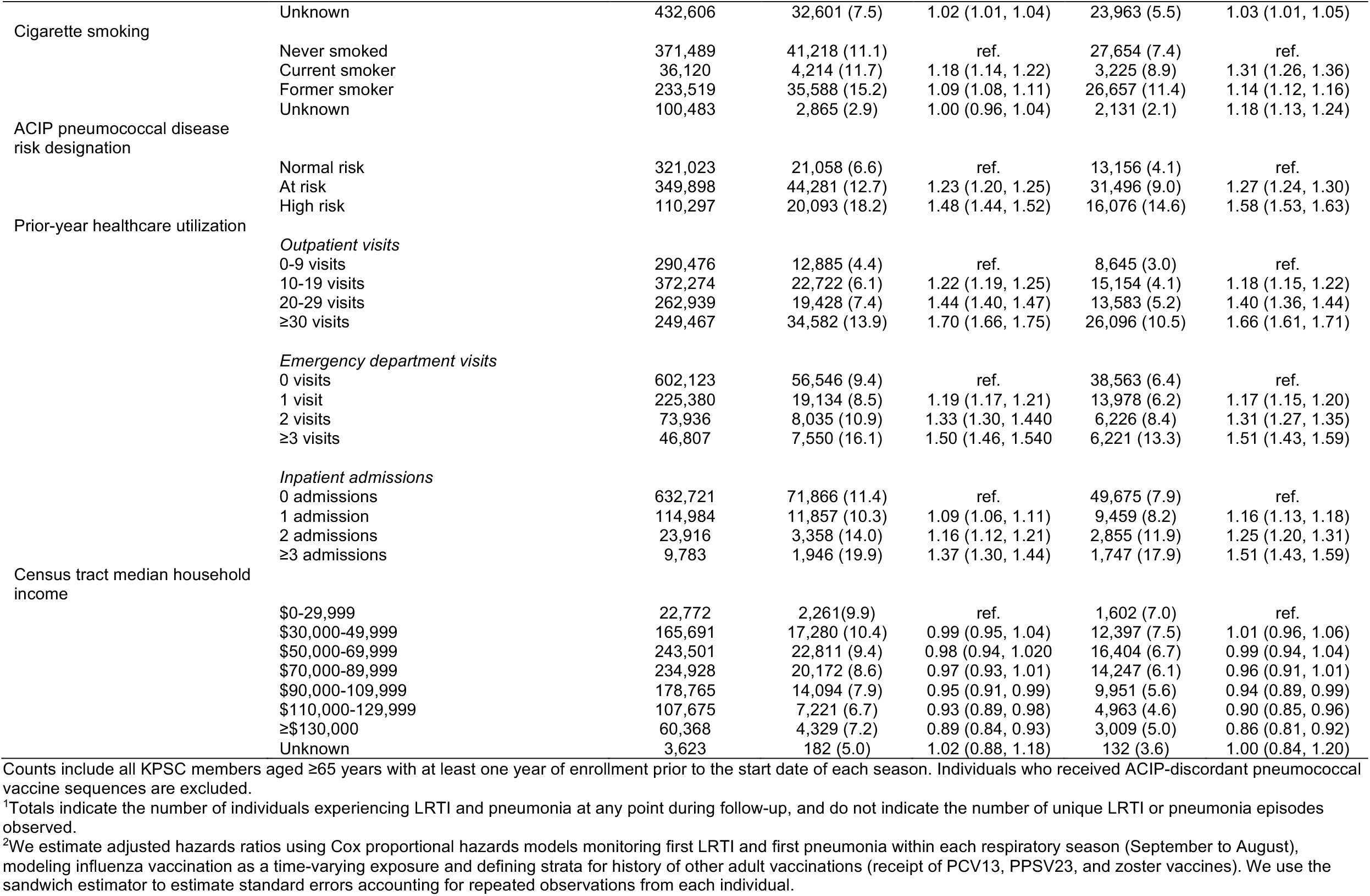
Risk factors for medically-attended lower respiratory tract infection and pneumonia due to any cause, among individuals aged ≥65 years within the full KPSC cohort.

**Table S9:** Incidence rates of medically-attended lower respiratory tract infection and pneumonia within the full KPSC cohort and analytic cohort, by history of receipt of each vaccine.

| Vaccine | Endpoint | Incidence per 100 person-years (first events/person-years at risk) |  |  |  |
| --- | --- | --- | --- | --- | --- |
|  |  | Full cohort (N=646,098) |  | Analytic cohort |  |
|  |  | <u>Vaccinated</u> | <u>Unvaccinated</u> | <u>Vaccinated</u> | <u>Unvaccinated</u> |
| PCV13 | LRTI | 6.9 (90,536/1,309,358) | 3.9 (8,195/210,703) | 5.6 (4,146/73,571) | 5.7 (1,456/25,764) |
|  | Pneumonia | 4.9 (64,199/1,323,313) | 2.7 (5,754/211,848) | 4.0 (2,950/74,187) | 3.8 (997/25,920) |
| PPSV23 | LRTI | 6.9 (93,908/1,359,177) | 3.0 (4,823/160,885) | 3.8 (1,245/32,932) | 3.8 (460/12,216) |
|  | Pneumonia | 4.9 (66,968/1,373,445) | 1.8 (2,985/161,717) | 2.2 (728/33,176) | 2.0 (245/12,285) |
| Seasonal influenza vaccine (inactivated or live attenuated)<sup>1</sup> | LRTI | 7.1 (63,244/893,521) | 5.6 (35,293/626,338) | 7.1 (63,244/893,521) | 6.3 (20,679/328,567) |
|  | Pneumonia | 4.8 (43,788/905,628) | 4.1 (25,992/629,331) | 4.8 (43,788/905,628) | 4.6 (15,350/330,362) |
| Any zoster vaccine | LRTI | 6.3 (42,046/672,641) | 6.7 (56,685/847,421) | 6.2 (1,188/19,207) | 5.3 (737/13,900) |
|  | Pneumonia | 4.2 (28,751/679,706) | 4.8 (41,202/855,456) | 4.3 (825/19,387) | 3.1 (442/14,049) |

Counts include all KPSC members aged ≥65 years with at least one year of enrollment prior to the start date of each season. Individuals who received ACIP-discordant pneumococcal vaccine sequences are excluded.
<sup>1</sup>All individuals receiving seasonal influenza vaccination are included in the analytic cohort for this exposure, as unvaccinated time at risk is observed for each individual who receives vaccination during the season.

**Table S10:** Effectiveness of PCV13 against medically-attended lower respiratory tract infection and pneumonia due to any cause, based on alternative respiratory season start dates.

| Season | Endpoint | Vaccine effectiveness (95% CI) |  |
| --- | --- | --- | --- |
|  |  | <u>Without risk factor adjustment</u> | <u>With risk factor adjustment</u> |
| July 1 to June 30 |  |  |  |
|  | LRTI | 7.1 (0.2, 13.5) | 6.3 (−0.7, 12.9) |
|  | Pneumonia | 7.9 (−0.3, 15.4) | 5.9 (−2.7, 13.8) |
| November 1 to October 31 |  |  |  |
|  | LRTI | 9.3 (1.7, 16.4) | 10.2 (2.5, 17.2) |
|  | Pneumonia | 11.3 (2.1, 19.6) | 11.7 (2.5, 20.0) |

Analyses follow the approach of those presented in **Figure 1**, defining years according to the start and end dates listed in the left-hand column.

**Table S11:** Effectiveness of other adult vaccines against medically-attended lower respiratory tract infection and pneumonia due to any cause.

| Vaccine | Endpoint | Vaccine effectiveness (95% CI) |  |
| --- | --- | --- | --- |
|  |  | <u>With adjustment for age and influenza vaccination (PPSV23 and zoster vaccine only)</u> | <u>With adjustment for age, influenza vaccination (PPSV23 and zoster vaccine only), and comorbid conditions</u> |
| PPSV23 | LRTI | -0.3 (-14.1, 11.9) | 3.9 (-9.4, 15.6) |
|  | Pneumonia | -3.9 (-23.7, 12.8) | 1.0 (-18.0, 17.0) |
| Seasonal influenza vaccine (inactivated or live attenuated)<sup>3,4</sup> | LRTI | 15.1 (13.1, 17.1) | 17.6 (15.6, 19.6) |
|  | Pneumonia | 16.3 (13.9, 18.7) | 18.0 (15.6, 20.3) |
| Any zoster vaccine | LRTI | -1.3 (-14.7, 10.6) | -1.7 (-15.3, 10.3) |
|  | Pneumonia | -2.0 (-19.0, 12.5) | -2.6 (-19.8, 12.1) |

**Table S12:** Seasonal influenza vaccine effectiveness against confirmed influenza A or B infection over the study period.

| Year | VE (95% CI), % |  |  |  |  |  | Source |
| --- | --- | --- | --- | --- | --- | --- | --- |
|  | All influenza A and B | All influenza A | Influenza A(H1N1)pdm09 | All influenza B | Influenza B/Yamagata | Influenza B/Victoria |  |
| 2016-17 | 40 (32, 46) | 33 (23, 41) | -- | -- | 52 (42, 61) | 56 (23, 75) | 1 |
| 2017-18 | 38 (31, 43) | 30 (22, 38) | 62 (50, 71) | -- | 48 (39, 55) | 76 (45, 89) | 2 |
| 2018-19 | 29 (21, 35) | -- | 44 (37, 51) | 44 (-12, 62) | -- | -- | 3 |
| 2019-20 | 39 (32, 44) |  | 30 (21, 39) |  |  | 45 (37, 52) | 4 |

Estimates of vaccine effectiveness are based on US-wide multi-site vaccine effectiveness surveillance data and not specific to the KPSC study population or setting.

